# Crowding and the epidemic intensity of COVID-19 transmission

**DOI:** 10.1101/2020.04.15.20064980

**Authors:** Benjamin Rader, Anjalika Nande, Ben Adlam, Alison L. Hill, Robert C. Reiner, David M. Pigott, Bernardo Gutierrez, COVID-19 data working group, John S. Brownstein, Marcia C. Castro, Huaiyu Tian, Oliver G. Pybus, Samuel V. Scarpino, Moritz UG Kraemer

## Abstract

The COVID-19 pandemic is straining public health systems worldwide and major non-pharmaceutical interventions have been implemented to slow its spread^1-4^. During the initial phase of the outbreak the spread was primarily determined by human mobility^5,6^. Yet empirical evidence on the effect of key geographic factors on local epidemic spread is lacking^7^. We analyse highly-resolved spatial variables for cities in China together with case count data in order to investigate the role of climate, urbanization, and variation in interventions across China. Here we show that the epidemic intensity of COVID-19 is strongly shaped by crowding, such that epidemics in dense cities are more spread out through time, and denser cities have larger total incidence. Observed differences in epidemic intensity are well captured by a metapopulation model of COVID-19 that explicitly accounts for spatial hierarchies. Densely-populated cities worldwide may experience more prolonged epidemics. Whilst stringent interventions can shorten the time length of these local epidemics, although these may be difficult to implement in many affected settings.

---

Predicting the epidemiology of the COVID-19 pandemic is a central priority for guiding epidemic responses around the world. China has undergone its first epidemic wave and, remarkably, cities across the country are now reporting few or no locally-acquired cases^8^. Analyses have indicated that that the spread of COVID-19 from Hubei to the rest of China was driven primarily by human mobility^6^ and the stringent measures to restrict human movement and public gatherings within and among cities in China have been associated with bringing local epidemics under control^5^. Key uncertainties remain as to which geographic factors drive local transmission dynamics and affect the intensity of transmission of COVID-19. For respiratory pathogens, “epidemic intensity” *(i.e*., the peakedness of the number of cases through time, or the shortest period during which the majority of cases are observed) varies with increased indoor crowding, and socio-economic and climatic factors^9–13^. Epidemic intensity is minimized when incidence is spread evenly across weeks and increases as incidence becomes more focused in particular days (**Figure 1C**, see a detailed description of how epidemic intensity is defined in Ref. ^9^). In any given location, higher epidemic intensity requires a larger surge capacity in the public health system^14^, especially for an emerging respiratory pathogen such as COVID-19^15^.

**Figure 1:**
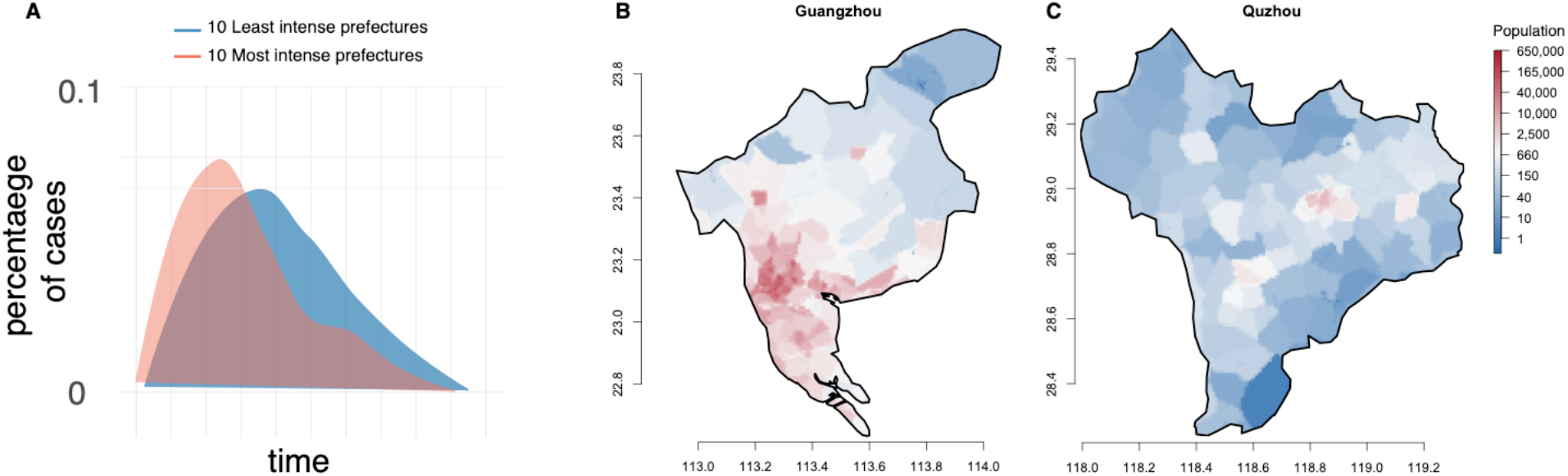
Maps of crowding in prefectures in China. A) shows epidemic curves that are normalized to show the percentage of cases that are occurring at each given day. The 10 most intense (red) prefectures are shown versus the 10 least intense (blue). B) An example of a prefecture with high levels of crowding (Guangzhou, Guangdong Province), versus (C) a prefecture where populations are more equally distributed across the prefecture (Quzhou, Zhejiang Province). The colour scale illustrates the number of inhabitants per grid cell (1km x 1km).

China provides richly detailed epidemiological time series^2,16,17^ across a wide range of geographic contexts, hence the epidemic there provides an opportunity to evaluate the role of factors in shaping the intensity of local epidemics. We use detailed line-list data from Chinese cities^18,19^, climate and population data, local human mobility data from Baidu, and timelines of outbreaks responses and interventions to identify drivers of local transmission in Chinese cities, with a focus on epidemic intensity among provinces in China.

To explore the impact of urbanization, temperature, and humidity, we used daily incidence data of confirmed COVID-19 cases (date of onset) aggregated at the prefectural level (n = 293) in China. Prefectures are administrative units that typically have one urban center (**Figure 1**). We aggregate individual level data that were collected from official government reports^17^. Epidemiological data in each prefecture were truncated to exclude dates before the first and after the last day of reported cases. The shape of epidemic curves varied between prefectures with some showing rapid rises and declines in cases and others showing more prolonged epidemics (**Figure 1A**). We estimate epidemic intensity for each prefecture from these data by calculating the inverse Shannon entropy of the distribution of incident cases^9^. We define the incidence distribution *p_ij_* for a given city to be the proportion of COVID-19 cases during epidemic wave *j* that occurred on day *i*. The inverse Shannon entropy of incidence for a given prefecture and year is then given by *ν_j_ =* (− *∑_i_p_ij_* log *p_ij_*)^−1^. Because *ν_j_* is a function of the disease incidence curve in each location, rather than of absolute incidence values, it is invariant under differences in overall reporting rates among cities or overall attack rates. Population counts for each prefecture were extracted from a 1 km x 1 km gridded surface of the world utilizing administrative-2 level cartographic boundaries.

Within each prefecture, we calculate Lloyd’s index of mean crowding^9,20^ treating the population count of each pixel as an individual unit (**Methods, Figure 1B and C**). The term ‘mean crowding’ used here is a specific metric that summarizes both, population density and how density is distributed across a prefecture (patchiness). Values on the resulting index above the mean pixel population count within each prefecture suggest a spatially-aggregated population structure (**Methods**). For example, Guangzhou has high values of crowding whilst Quzhou which has a more evenly distributed population in its prefecture (**Figure 1B and C**). Using the centroid of each prefecture we calculate daily mean temperature and specific humidity; these values are subsequently averaged over each prefecture’s reporting period (**Methods**). We performed log-linear regression modeling to determine the association between epidemic intensity with the socio-economic and environmental variables (**Methods**).

We found that epidemic intensity is significantly negatively correlated with mean population crowding and varies widely across the country (**Figure 2**, Extended Data Table 1, p-value < 0.001). Our observation contrasts those expected from simple and classical epidemiological models where it would be expected to see more intensity in crowded areas^21,22^. We hypothesize that the mechanism that underlies the more crowded cities experience less intense outbreaks because crowding enables more widespread and sustained transmission between households leading incidence to be more widely distributed in time (see section below for detailed simulation, **Methods**). Population size, mean temperature, and mean specific humidity were all significant but their correlation coefficients were much smaller (**Extended Data Table 1**). A multivariate-model was able to explain a large fraction of the variation in epidemic intensity across Chinese cities (R^2^ = 0.54). We perform sensitivity analysis to account for potential noise in the city level incidence distribution (Extended Data Fig. 1).

**Figure 2.**
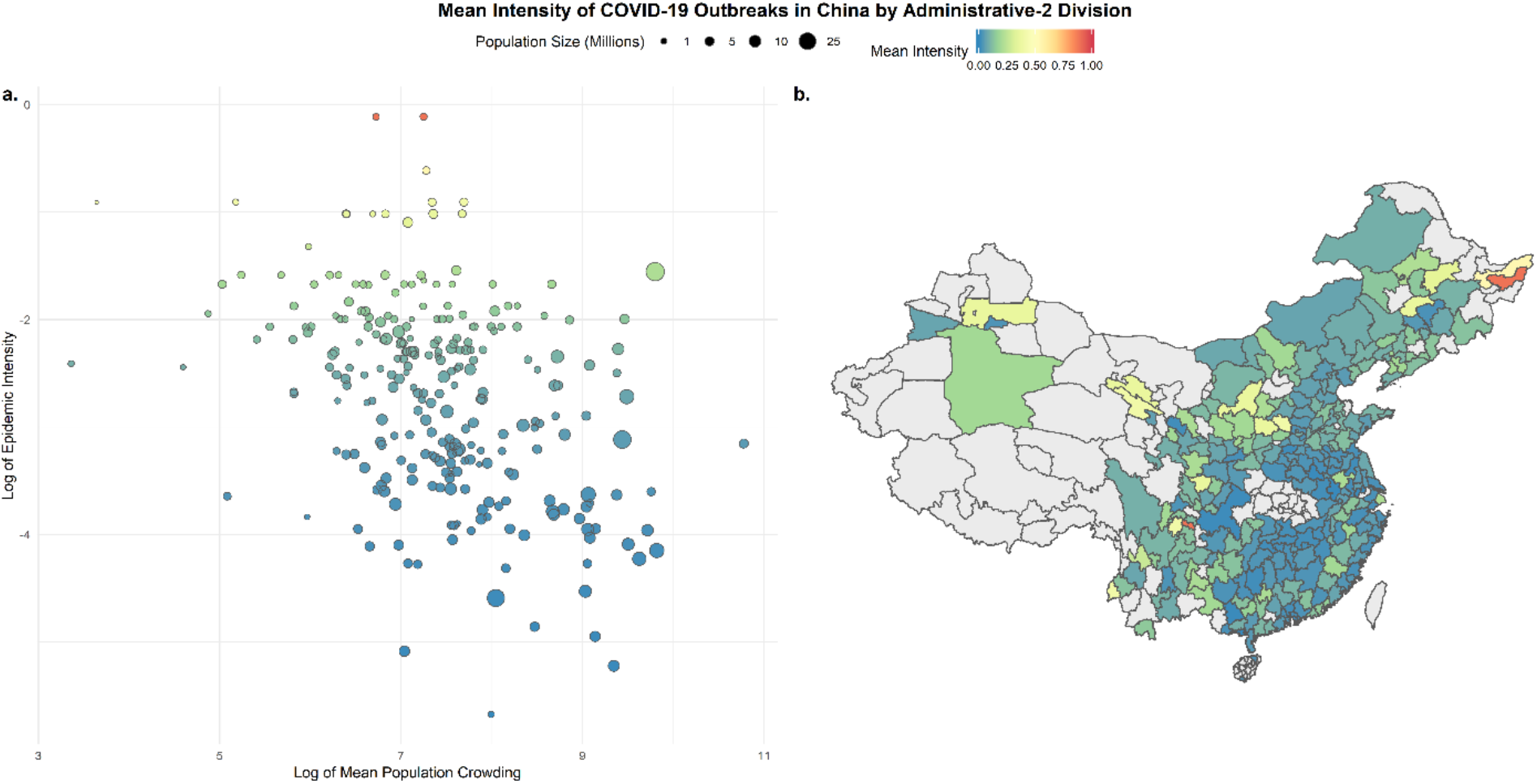
Crowding and the intensity of transmission of COVID-19 in China. a) negative association between log of epidemic intensity, as measured by inverse Shannon entropy (Methods), and log population crowding, as measure by Lloyd’s mean crowding (Methods). Lower intensity and therefore prolonged epidemics in larger cities. The size of the points indicate the size of the population in each city, b) Map of epidemic intensity in China at the prefecture level. Darker colours indicate lower intensity and lighter colours higher intensity. Grey prefectures had not enough reported cases, no cases or were not included in analyses (Hubei Province).

One key uncertainty in previous applications of models of epidemic intensity was the contribution of disease importation(s) on the shape of the epidemic^9^. Due to the unprecedented scale of human mobility restrictions imposed in China, the fact that the early epidemic was effectively from a single source, coupled with the availability of real-time data on mobility, we can evaluate the impact of these restrictions on the epidemic intensity relative to the local dynamics. To do so, we performed a univariate analysis (**Extended Data Table 1**) and found that human mobility explained 14% of the variation in epidemic intensity. This further supports earlier findings that COVID-19 had already spread throughout much of China prior to the cordon sanitaire of Hubei province and that the pattern of seeding potentially modulates epidemic intensity^6,23^. These findings are in agreement with previous work on other pathogens (measles, influenza) which showed that once local epidemics are established case importation becomes less important in determining epidemic intensity^24^.

To evaluate the potential impact of variability of intensity on the peak incidence and total incidence we performed a simple linear regression. We found that peak incidence was correlated with epidemic intensity (locations that had high intensity also had more cases at the peak). Total incidence, however, was larger in areas with lower estimated intensity, which is intuitive as crowded areas have longer epidemics that affect more people (**Extended Data Table 2**). This suggests that measures taken to mitigate the epidemic may need to be enforced more strictly in smaller cities to lower the peak incidence (flatten the curve) but conversely may not need to be implemented as long. Furthermore, with lower total incidence in small cities, the risk of resurgence may be elevated due to lower population immunity. There is urgent need to collect serological evidence to provide a full picture of attack rates across the world^25^.

Using our model trained on cities in China we extrapolated epidemic intensity to cities across the world (**Figure 3**). Figure 3 shows the distribution of epidemic intensity in 380 urban centers. Cities in yellow are predicted to have higher epidemic intensity relative to those in blue (a full list is provided in **Extended Data Table 3**). Small inland cities in sub-Saharan Africa had high predicted epidemic intensity and may be particularly prone to experience large surge capacity in the public health system^26^. In general, coastal cities had lower predicted intensity and larger and more prolonged predicted epidemics. Global predictions of epidemic intensity in cities rely on fitted relationships of the first epidemic curve from Chinese prefectures and therefore need to be interpreted with extreme caution.

**Figure 3:**
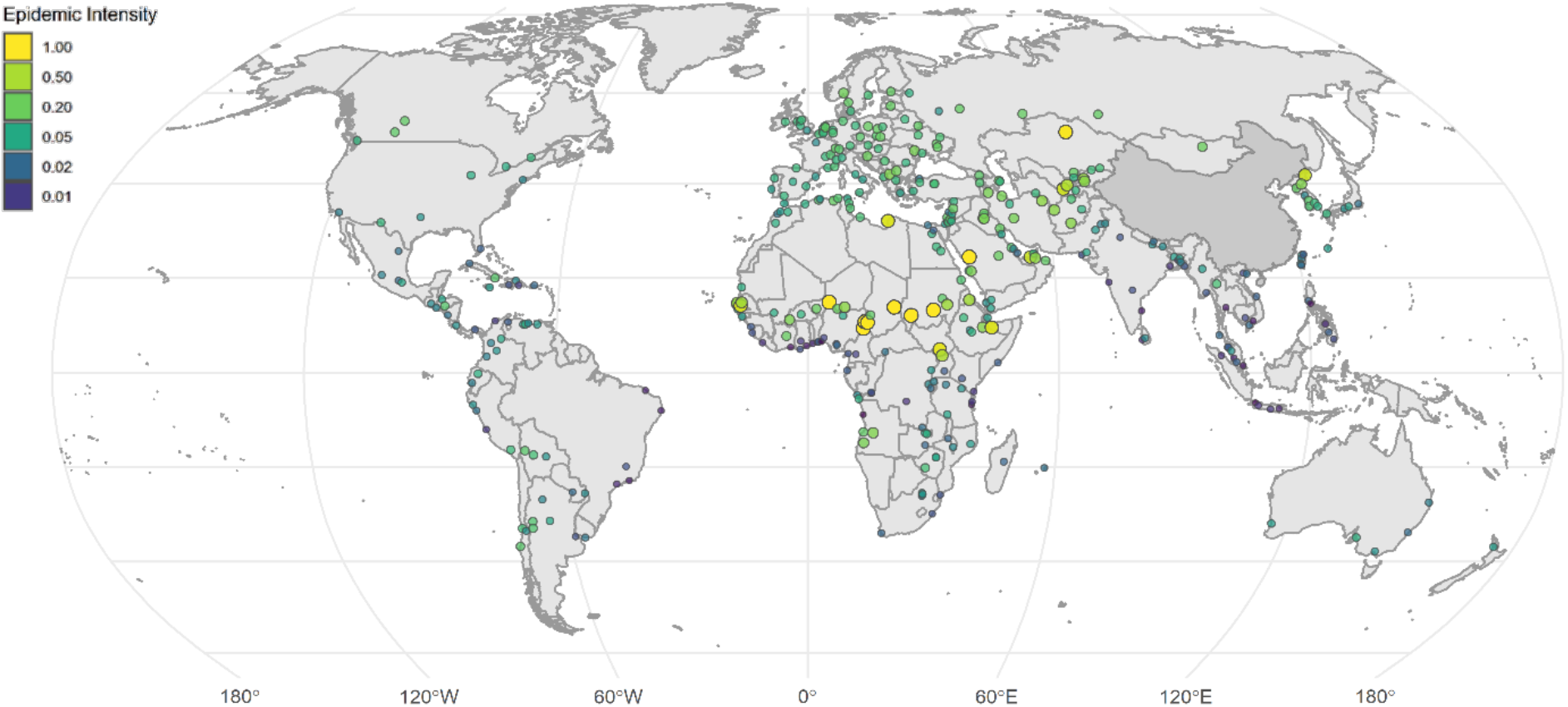
Predicted epidemic intensities vary across 380 global cities. Darker colours represent low epidemic intensity and lighter colours represent high epidemic intensity. Estimates were generated using the full model (Model 5) fitted to epidemic curves in Chinese cities (Extended Data Table 1). A full list of epidemic intensities can be found in the Extended Data Table 3. Epidemic intensity is a measure of peakedness of epidemics and does not reflect the expected number of cases (Methods).

To understand the mechanism responsible for our finding that outbreaks in crowded cities were lower intensity *(i.e*. more spread out in time), we simulated stochastic epidemic dynamics in different types of populations. Simple, well-mixed transmission models where contact rates are higher in crowded regions were not consistent with our findings, since they predict crowded regions would have more intense and higher-peaked outbreaks. To capture more realistic contact patterns, we created hierarchically-structured populations^27^ where individuals had high rates of contact within their households (households are defined broadly and could represent care homes, hospitals, prisons, etc.), lower rates with individuals from other households but within the same “neighborhoods”, and relatively rare contact with other individuals in the same prefecture (**Figure 4A**). Assumptions are consistent with reports that the majority of onward transmission occurred in households^2,28^. We assumed spread between prefectures was negligible once an outbreak started. In this scenario, “sparse” prefectures often had more intense, short-term outbreaks that were isolated to certain neighborhoods, while “crowded” prefectures could have drawn-out, low intensity outbreaks that jumped between the more highly-connected “neighborhoods” (**Figures 4B and C**). These outbreaks had larger final size than those in less-crowded areas (**Figure 4C**) which likely is related to large overdispersion in the reproduction number of COVID-19^29,30^ where local outbreaks can reach their full potential due to the availability of contacts. We also considered outbreak dynamics in sparse and crowded prefectures under strong social distancing measures, which is likely to be the scenario occurring across China during most of the time captured by our study and certainly after January 23, 2020^2^. If social distancing reduces non-household contacts by the same relative amount in all prefectures, there will be more contacts remaining in crowded areas, since baseline contact rates are higher. In this case, it may take much longer for the infection to die out post-intervention in crowded areas (**Figure 4D**), leading to a lower intensity outbreak with larger final size, as seen in our data (**Figure 1C**).

**Figure 4:**
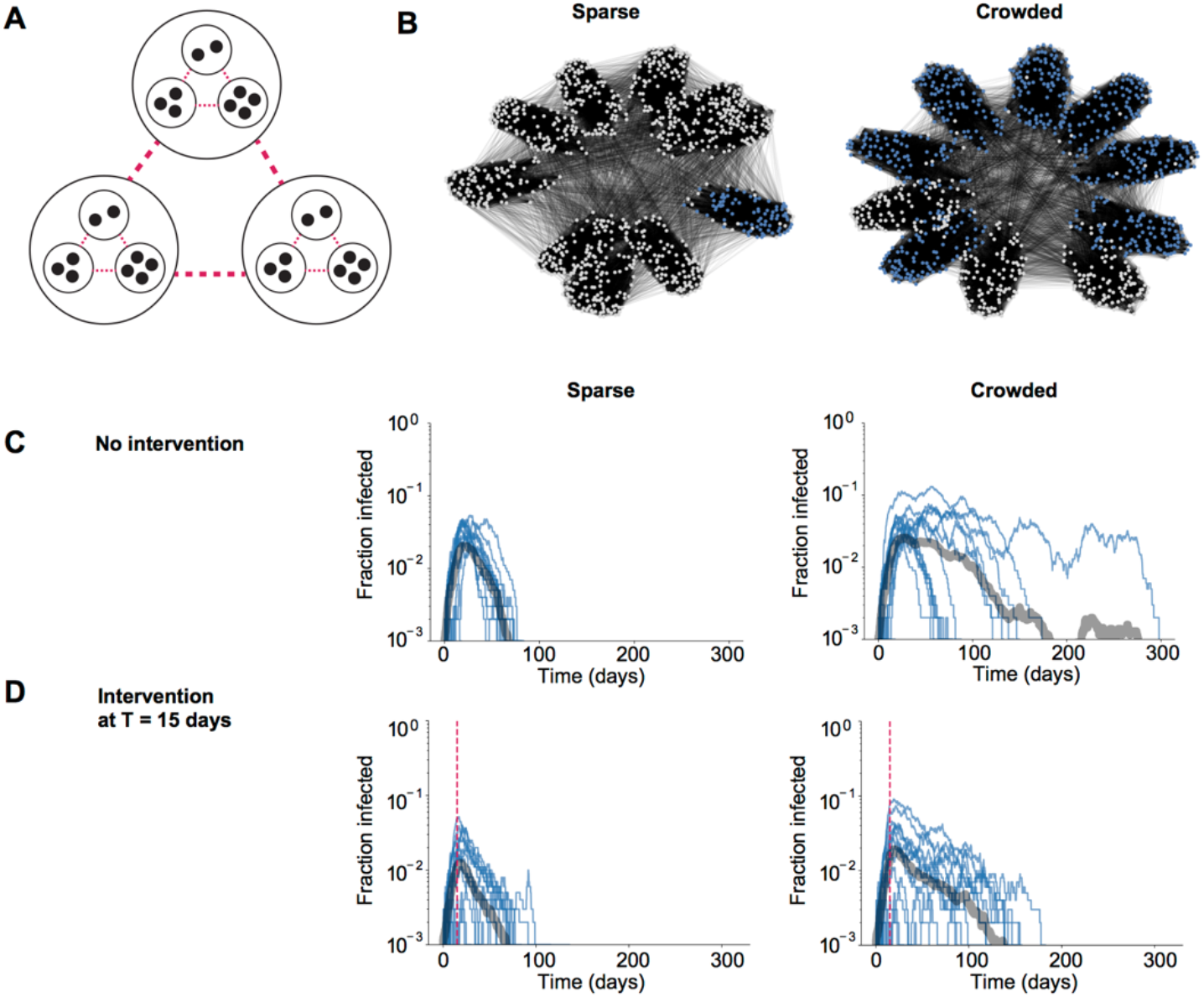
Mechanisms generating less intense epidemics in crowded populations. A) Schematic of a hierarchically-structured population model consisting of households and “neighborhoods” within a prefecture. Transmission is more likely among contacts connected at lower spatial levels. Crowded populations have stronger connections outside the household, and interventions reduce the strength of these connections in both populations (pink lines). B) - C) Simulated outbreak dynamics in the absence of interventions in crowded vs sparse populations. For the networks in (B), blue nodes are individuals who were eventually infected by the end of the outbreak. In (C), individual realizations are shown with thin blue lines and the average in the thick grey line. D) Simulated outbreak dynamics in the presence of strong social distancing measures in crowded vs. sparse populations. The intervention was implemented at day 15 (pink line) and led to a 75% reduction in contacts.

Spatial covariates and particularly crowding are important parameters to consider in the assessment of epidemics across the world. Crowded cities tend to be more prolonged due to increased crowding and the higher potential for transmission chains to persist *(i.e*., in denser environments there is higher potential for two randomly selected hosts in a population to attain spatiotemporal proximity sufficient for COVID-19 transmission). Our findings confirm previous work on epidemic intensity of transmission of influenza in cities^9^ albeit the mechanism for influenza is likely driven by the accumulation of immunity rather than the specific network structure of individuals. More generally, our work provides empirical support for the role of spatial organization in determining infectious disease dynamics and the limited capacity of *cordon sanitaires* to control local epidemics^27,31^. We were unable to test more specific hypotheses about which interventions may have impacted the intensity of transmission within and between cities. Further, even though humidity was negatively associated with epidemic intensity it did not explain the majority of the variation and more work will be needed to find causal evidence for the effect of humidity on transmission dynamics of COVID-19. Therefore, maps showing epidemic intensity in cities outside China (**Figure 3**) should be interpreted with particular caution.

Currently, non-pharmaceutical interventions are the primary control strategy for COVID-19. As a result, public health measures are often focused on ‘flattening the curve’ to lower the risk of essential services running out of capacity. We show that spatial context, especially crowding, can result in a higher risk of intensive epidemics in less crowded, comparatively rural areas. Therefore, it will be critical to view non-pharmaceutical interventions through the perspective of crowding *(i.e*., how does an intervention reduce the circle of contacts of an average individual) in cities across the world. Specifically, cities in sub-Saharan Africa have high predicted epidemic intensities that will likely overwhelm already stressed health care systems.

## Data Availability

Data availability: We collated epidemiological data from publicly available data sources (news articles, press releases and published reports from public health agencies) which are described in full here17. All the epidemiological information that we used is documented in the main text, the extended data, and supplementary tables.

https://www.nature.com/articles/s41597-020-0448-0

## Methods

### Epidemiological data

No officially reported line list was available for cases in China. We use a standardised protocol^32^ to extract individual level data from December 1st, 2019 - March 30^th^, 2020. Sources are mainly official reports from provincial, municipal or national health governments. Data included basic demographics (age, sex), travel histories and key dates (dates of onset of symptoms, hospitalization, and confirmation). Data were entered by a team of data curators on a rolling basis and technical validation and geo-positioning protocols were applied continuously to ensure validity. A detailed description of the methodology is available^17^. Lastly, total numbers were matched with officially reported data from China and other government reports.

### Estimating epidemic intensity

Epidemic intensity was estimated for each prefecture by calculating the inverse Shannon entropy of the distribution of COVID-19 cases. Shannon entropy was used to fit time series of other respiratory infections (influenza)^9^. The Shannon entropy of incidence for a given prefecture and year is then given by *ν_j_ =* (− ∑*_i_p_ij_* log *p_ij_*)^−1^. Because *ν_j_* is a function of incidence distribution in each location rather than raw incidence it is invariant under differences in overall reporting rates between cities or attack rates. We then assessed how mean intensity *ν ∝ ∑_j_ν_j_* varied across geographic areas in China.

### Proxies for COVID-19 interventions

Real-time measures of human mobility were extracted from the Baidu Qianxi web platform to estimate the proportion of daily movement between the city of Wuhan to Hubei and 30 other Chinese provinces. Relative mobility volume was available from January 2, 2020 to January 25, 2020 and averaged across these dates to create a single measure of relative flows from Wuhan. This data was only available at the province level, so each individual prefecture inherited the relative mobility of its higher-level province. Baidu’s mapping service is estimated to have a 30% market share in China and more data can be found^5,6^.

### Data on drivers of transmission of COVID-19

Prefecture-specific population counts and densities were derived from the 2020 Gridded Population of The World, a modeled continuous surface of population estimated from national census data and the United Nations World Population Prospectus^33^. Population counts are defined at a 30 arc-second resolution (approximately 1 km x 1 km at the equator) and extracted within administrative-2 level cartographic boundaries defined by the National Bureau of Statistics of China. Lloyd’s mean crowding, 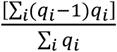, was estimated for each prefecture where *q_i_* represents the population count of each non-zero pixel within a prefecture’s boundary and the resulting value estimates an individual’s mean number of expected neighbors^9,34^.

Daily temperature (°F), relative humidity (%) and atmospheric pressure (Pa) at the centroid of each prefecture was provided by The Dark Sky Company via the Dark Sky API and aggregated across a variety of data sources. Specific humidity (kg/kg) was then calculated using the R package, humidity^12^. Meteorological variables for each prefecture were then averaged across the entirety of the study period.

### Statistical analysis

We normalized the values of epidemic intensity between 0 and 1, and for all non-zero values fit a Generalized Linear Model (GLM) of the form:

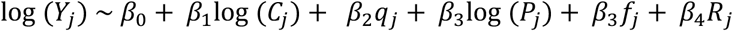

where for each prefecture *j, Y* is the scaled Shannon-diversity measure of epidemic intensity derived from the COVID-19 time series, *C* is Lloyd’s Index of Mean Crowding^20,35^, *q* is the mean specific humidity over the reporting period in kg/kg, *P* is the estimated population count and *f* is the relative population flows from Wuhan to each prefecture’s higher level province. To account for the length of the epidemic period in each city we calculate *R* as the number of reporting days.

### Projecting epidemic intensity in cities around the world

We selected 380 urban centers from the European Commission Global Human Settlement Urban Centre Database and their included cartographic boundaries^36^. To ensure global coverage, up to the five most populous cities in each country were selected from the 1,000 most populous urban centers recorded in the database. Population count, crowding, and meteorological variables were then estimated following identical procedures used to calculate these variables in the Chinese prefectures. Weather measurements were averaged over the 2-month period starting on February 1, 2020.

The parameters from the model of epidemic intensity predicted by humidity, crowding and population size (see Table 1, Model 6) were used to estimate relative intensity in the 380 urban centers. Predicted values of epidemic intensity that fell outside the original covariate space [0,1] (n=7) were set to 1. A full list of predicted epidemic intensities can be found in the Supplementary Information.

### Sensitivity analyses

The inverse Shannon entropy metric may be sensitive to noise in incidence distribution. For example, the noisier the incidence distribution the higher the epidemic intensity. To the extent that noise is elevated in small populations (due to demographic stochasticity for instance) intensity also tends to be higher in smaller populations, even if they have the same underlying shape to their epidemic curve. Lloyd’s mean crowding also varies strongly with population size. Therefore, some of the observed relationship between intensity and crowding may be due to (possibly independent) statistical scaling of both intensity and crowding with population size. We therefore perform sensitivity analysis to test if cities that are more crowded than expected for their size have lower epidemic intensities than expected for their size. We calculate ‘excess intensity’ as the residuals on a regression of log(epidemic intensity) ~ log(pop); ‘excess crowing’ as the residuals on a regression of log(crowding) ~ log(pop) and plot the relationship between excess intensity and excess crowding’ (Extended Data Figure 1).

### Simulating epidemic dynamics

We simulated a simple stochastic SIR model of infection spread on weighted networks created to represent hierarchically-structured populations. Individuals were first assigned to households using the distribution of household sizes in China (data from UN Population Division, mean 3.4 individuals). Households were then assigned to “neighborhoods” of ~100 individuals, and all neighborhood members were connected with a lower weight. A randomly-chosen 10% of individuals were given “external” connections to individuals outside the neighborhood. The total population size was N=1000. Simulations were run for 300 days and averages were taken over 20 iterations. The SIR model used a per-contact transmission rate of *β*=0.15/day and recovery rate *γ*=0.1/day. For the simulations without interventions, the weights were *w_HH_* = 1, *w_NH_* = 0.01, and *w_EX_* = 0.001 for the “crowded” prefecture and *w_EX_* = 0.0001 for the “sparse” prefecture. For the simulations with interventions, the household and neighborhood weights were the same but we used *w_EX_* = 0.01 for the “crowded” prefecture and *w_EX_* = 0.001 for the “sparse” prefecture. The intervention reduced the weight of all connections outside the household by 75%.

## Acknowledgements

The authors thank Kathryn Cordiano for her statistical assistance. BR acknowledges funding from Google.org. MUGK acknowledges funding from European Commission H2020 program (MOOD project) and a Branco Weiss Fellowship. OGP and HT acknowledge funding from the Oxford Martin School. ALH and AN acknowledge funding from the US National Institutes of Health (DP5OD019851). The funding bodies had no role in study design, data collection and analysis, preparation of the manuscript, or the decision to publish. All authors have seen and approved the manuscript.

## Author contributions

MUGK, SVS, OGP conceived the research. BR, ALH, AN, BA, SVS, MUGK analysed the data. MUGK wrote the first draft of the manuscript. All authors contributed to interpretation of results and manuscript writing.

## Data availability

We collated epidemiological data from publicly available data sources (news articles, press releases and published reports from public health agencies) which are described in full here^17^. All the epidemiological information that we used is documented in the main text, the extended data, and supplementary tables.

## Code availability

The code is available from this link: tbc and the simulation code is available from here: https://github.com/alsnhll/SIRNestedNetwork

